# In-line chlorination for drinking water in rural Odisha, India: a randomized controlled implementation trial

**DOI:** 10.1101/2025.07.10.25331308

**Authors:** Jeremy Lowe, Vaishnavi Prathap, Akito Kamei, Sidhartha Giri, Krushna Chandra Sahoo, Michael Kremer, Elisa M. Maffioli, Amy J. Pickering

## Abstract

In-line chlorination automatically treats drinking water in piped water systems or at points-of-collection, substantially reducing morbidity and mortality while averting the burden of water treatment on individuals. Approximately 2.3 billion people globally use fecally contaminated drinking water infrastructure that is potentially compatible with in-line chlorination. In India, the Ministry of Jal Shakti’s Jal Jeevan Mission has increased access to piped drinking water among rural households, however, water is often supplied intermittently without treatment. The aims of this study were to 1) test installation and operational procedures for in-line chlorination in rural drinking water systems, 2) evaluate acceptability and adoption of chlorinated water, and 3) assess reductions in drinking water fecal contamination, including with antibiotic resistant bacteria. We conducted a randomized controlled implementation trial of in-line chlorination in 20 villages in Rayagada district, Odisha, India over a 1-year period. Data collection included a baseline and post-intervention census (N=914) and surveys with a random subset of households over six timepoints (N=1,041) to assess water quality. We discuss operational challenges, including maintaining consistent chlorine dosing and addressing taste and odor preferences. In the treatment group, we detected free chlorine residual in 51% of tap water samples; this prevalence rose to 80% after increasing the target dose. Despite taste complaints in treatment communities, participants continued using piped water as their primary drinking water source. Individuals in treatment communities reported reduced time spent manually treating their drinking water, a responsibility borne by women in 92% of households. Treatment reduced *E. coli* prevalence by 70% in household tap water and by 47% in stored water, as well as reduced the presence of antibiotic resistant *E. coli*. In-line chlorination can lead to better quality water, lower the time burden of water treatment, and increase user adoption of chlorinated drinking water, but achieving this requires dedicated implementation and monitoring.

## Introduction

Sustainable Development Goal (SDG) 6.1 aims to provide universal access to safely managed drinking water by 2030, which is defined as greater than 99% of the population having access to an improved water source that is accessible on premises, available when needed, and free from fecal and priority chemical contamination (1). However, current rates of global progress need to be quadrupled to meet SDG 6.1 by 2030 (1). New evidence suggests that 1 in 3 people in low- and middle-income countries (LMICs) lack access to safely managed drinking water (2). Fecal contamination is the limiting factor in achieving access to safely managed drinking water for almost half of the population living in LMICs (2), emphasizing the importance of water treatment in achieving universal access to safe drinking water.

Drinking water treatment is a cost-effective strategy to reduce diarrheal disease and child mortality globally. A systematic review found that water treatment interventions can reduce diarrhea in children under five by about 24% as compared to settings without water treatment (3). A recent meta-analysis of 18 water treatment randomized controlled trials (RCTs) also suggests a 24% reduction in all-cause mortality in children under five from water treatment, primarily driven by the use of chlorination, and a cost per disability-adjusted life year (DALY) averted of about USD 40 (4). The transmission of antibiotic resistance in the environment is also widely recognized as a threat to public health (5–8), yet no RCTs have evaluated the impact that improved water treatment infrastructure can have on reducing human exposure to antibiotic resistance through drinking water (7,9).

Achieving high and sustained adoption rates is a key challenge for water treatment interventions. Point-of-use water treatment methods such as boiling or manual chlorination require sustained behavior change, the burden of which often falls on women and often leads to lower adoption A systematic review finds average adoption rates of 47% for point-of-use chlorination, falling to 11% for studies in which participants were visited less than once a month by study staff (10). Some studies have found reductions in treatment after interventions or regular visits ended (11). Perhaps due to low adoption, some studies of point-of-use water treatment find null effects on child health outcomes (12–15). Other evidence suggests that consumers are often willing to pay for convenient access to water but have low willingness to pay for water treatment (16), making it challenging to sustain programs through user fees for water treatment.

In-line chlorination is a promising low-cost water treatment solution that can address implementation barriers to scale (17). Chlorination has been historically used to disinfect drinking water supplies and has reduced enteric disease burden and mortality globally (18). In-line chlorination is a drinking water treatment method that reduces cost and time burdens on individuals by automatically chlorinating water from community-level water points without requiring electricity or daily operation. A variety of in-line chlorination technologies are now available in the market, including open source products such as chlorine tablet erosion dosers that can be constructed from locally-available plumbing materials (17). A previous RCT in Dhaka, Bangladesh, observed a 23% reduction in child diarrhea following an in-line chlorination intervention in community water systems, with a detectable free chlorine residual at household tap connections 83% of the time (19). Approximately 2.3 billion individuals globally use drinking water infrastructure that is potentially compatible with in-line chlorination, presenting an opportunity to continue scaling up across multiple contexts (20). However, before in-line chlorination can be implemented at such a large scale, effective operational procedures need to be evaluated to support implementers across diverse infrastructure and institutional contexts.

In India, the Ministry of Jal Shakti’s Jal Jeevan Mission aimed to supply safe drinking water through household tap connections to all rural Indian households by 2024 and was recently extended to 2028 (21,22). As of December 2024, 79.4% of households in India had access to a tap connection providing at least 55 liters per capita per day (23). The Jal Jeevan Mission supported state governments in the construction of new drinking water supply infrastructure and retrofitting old water supply schemes to achieve the large increase in household piped water access (24). Through the Jal Jeevan Mission, drinking water is supplied to rural villages either through single-village or multi-village distribution networks in which water is pumped from boreholes to elevated storage tanks and gravity-fed through pipes to household tap connections (24). However, water is only supplied intermittently for a few hours each day, which can introduce microbiological contamination (25,26). Intermittent water supply is estimated to cause about 109,000 diarrhea-related DALYs per year globally (27). In-line chlorination is a potentially compatible and scalable treatment strategy for drinking water infrastructure delivered through the Jal Jeevan Mission. However, in single-village schemes (which serve approximately 60% of rural India), there is typically no system for automatic chlorination, thus requiring daily manual effort by village pump operators to conduct water treatment. The effectiveness of in-line chlorination in providing sustained water treatment in this context is relatively unknown.

We conducted a randomized controlled implementation trial of in-line chlorination in rural Indian villages served by the Jal Jeevan Mission. The aim of this study was to field test installation and operation procedures to inform local guidelines for implementing in-line chlorination and understand if chlorinated water would be acceptable to users. In addition, we evaluated how effectively in-line chlorination could chlorinate drinking water and its impact on reducing levels of fecal contamination and antibiotic resistant fecal bacteria. This study was also conducted to assess the feasibility of a larger study powered to detect the effects of in-line chlorination on health outcomes.

## Materials and Methods

### Study Design

We conducted a cluster-randomized controlled implementation trial in 20 villages in rural Odisha, India; 10 villages were randomly assigned to receive an installation of in-line chlorine dosers and 10 to a control group where no intervention took place. Villages were enrolled into the study following an infrastructure assessment by a civil engineer to determine if the piped water infrastructure was compatible with in-line chlorination.

Villages were considered eligible for the study if they met the following criteria: 1) the piped water infrastructure supplied water to a single village through a central tank and was compatible with a tablet-based chlorine doser at the inlet or outlet of the tank, 2) the turbidity of drinking water sources was less than 5 nephelometric turbidity units, 3) the water distribution infrastructure did not have any major issues preventing water supply or in-line chlorination installation (e.g. major leaks at valves, dysfunctional borewell pump, major disruptions in electricity supply lasting multiple days, etc.), 4) water was supplied at least once daily for any amount of time through household tap connections, and 5) there were no plans for the village piped water supply to be converted to a larger distribution scheme connected to a centralized drinking water treatment plant in the following year.

Based on administrative information shared by the Department of Panchayati Raj and Drinking Water, we selected Rayagada district of Odisha given the relatively high prevalence of single-village water distribution schemes compared to other districts. In consultation with the department, we identified villages reporting near complete coverage of tap connections in households and served by a single-village water distribution scheme. Among these, a civil engineer visited villages to assess the compatibility of the piped water supply infrastructure with in-line chlorination (see above criteria). The civil engineer visited 33 villages in total and identified 22 eligible villages in Rayagada for the study in August 2023. 20 villages were enrolled in the study, while an additional two villages were only enrolled at baseline as backup villages in the event a village needed to be replaced for major infrastructure failures (i.e. pipeline breaks). The sample size was limited by the study budget. After baseline data collection but before intervention rollout, one of the treatment villages had to be dropped and replaced with a backup village from another block due to a breakage in the supply pipeline resulting in people primarily using alternative water sources besides their household tap connections.

Before the in-line chlorination implementation, we conducted a baseline census where all households were visited to identify households with mothers and children under five (N = 914) in 20 villages. A subset of households (N = 200) was assessed for water use practices and water quality, including measurements for free and total chlorine and *Escherichia coli* (*E. coli*). Villages were then randomly assigned to control or treatment groups, where 10 out of 20 villages were randomly assigned to receive the in-line chlorination intervention. The randomization was stratified by block (a sub-district administrative unit) and if the village was the Gram Panchayat headquarters. Among the treatment group, we implemented two in-line chlorination technologies that automatically dose chlorine into piped water supply without the need for electricity via chlorine tablet erosion: the PurAll 100 (28) and an open-source technology called CTI-8 (29). We followed up in the 20 villages over 1 year through 6 data collection rounds to assess changes in water use practices and water quality (Figure 1).

**Figure 1:**
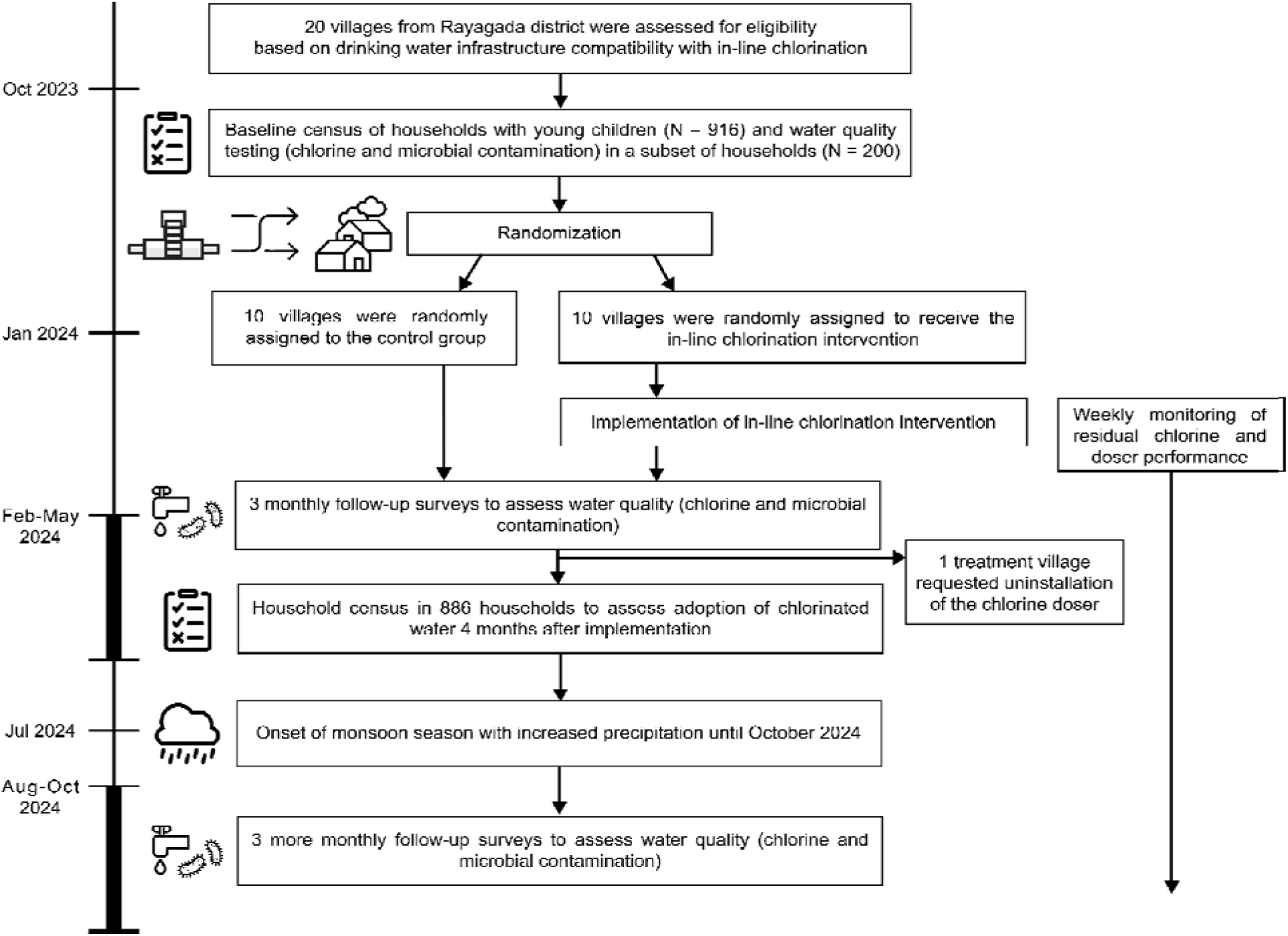
Timeline of study activities.

The study was conducted in collaboration with the Department of Drinking Water and Sanitation, Ministry of Jal Shakti, Government of India, and the Department of Panchayati Raj and Drinking Water, Government of Odisha. In August 2023, the Government of Odisha issued a permission letter to facilitate the RCT in villages in Rayagada district. Research ethics approvals were received from the Institutional Ethics Committee of ICMR-Regional Medical Research Centres, Bhubaneswar. This study was also approved by the Directorate of Health Services, Government of Odisha. Institutional review board approval for human subjects research was received from the University of Chicago (IRB 23-0842) and the Institute for Financial Management and Research in India (IRB #10862).

### Intervention delivery

We used two chlorine tablet-based in-line chlorination technologies in the intervention: the commercially available PurAll 100 (28) and open-source CTI-8 (29) (SI Figure 1). Each tablet doser operated by redirecting a proportion of flow through a cartridge containing solid trichloroisocyanuric acid (TCCA) tablets, which would passively dissolve and dose chlorine into the drinking water pipeline via a bypass line. We used the PurAll 100 based on its commercial availability in India and the open-source CTI-8 because it could be assembled on-site using locally available materials. Both technologies were also compatible with existing drinking water infrastructure. Drinking water in the study villages was supplied intermittently by pumping groundwater into an elevated storage tank (40,000-100,000 L capacity) via an electric borehole pump; water was then released by gravity flow through the distribution network to individual household tap connections once or twice per day for one to two hours on average by the pump operator. Villages ranged in size between 67 and 380 households, and each household had their own tap connection. We installed each tablet doser on the inlet pipeline leading into the tank which allowed chlorinated water to be disinfected in the storage tank between supply cycles. The material costs for the parts to build the CTI-8 device were approximately 4000 INR ($47 USD), and the PurAll 100 device cost approximately 25500 INR ($307 USD). A PurAll 100 chlorine tablet cartridge used ten 200 gram TCCA tablets and cost ∼ 4700 INR ($57 USD) and was advertised to treat 2.5 million liters of water at 1 mg/L concentration. The equal-capacity CTI-8 chlorine tablet refill cartridge cost ∼ 432 INR ($5 USD) with tablets purchased from an alternative vendor (Acuro Organics Limited, New Delhi, India). In our study, we observed the cost per 1000 liters of treated water with a free chlorine residual of 0.2-0.6 mg/L was about $0.04 per 1000 liters for the PurAll chlorine tablets and $0.004 per 1000 liters for the CTI-8 with chlorine tablets from the alternative vendor (based on the average volume of water supplied per day, 64,000 liters, and the average number of days between refills, 21.4 days).

Gram Vikas, a non-governmental organization operating throughout Odisha, co-designed the intervention and led the implementation. Prior to installation of the in-line chlorination technologies, Gram Vikas conducted two community sensitization meetings. The first meeting discussed the purpose of in-line chlorination with the village sarpanch, a locally elected leader to the Gram Panchayat, and borehole pump operator; this was followed by a village meeting with general community members to introduce in-line chlorination as a water treatment method and answer questions. Additional meetings were conducted on an as-needed basis by a social coordinator and resulted in approximately two to four more village meetings per community over the study period. Between December 2023 and January 2024, PurAll 100 tablet dosers were installed in 4 villages, and CTI-8 tablet dosers were installed in 6 villages. Tablet dosers are often designed to operate under low pressure or unpressurized systems, but we successfully adapted these to operate under pressurized conditions at ground level before water was piped into elevated storage tanks. In 3 villages, tanks had two inlet lines (e.g. two borewells pumping into the tank) and we installed one device on each tank inlet line. In one village, an inlet installation was not feasible because the entire inlet line was routed under a concrete platform, but we were able to achieve effective dosing by installing two tablet dosers in series on the outlet line. A plumber and engineer visited each site twice: once to gather measurements for required materials and a second time to complete the installation which took between 1 to 4 hours.

We targeted a free chlorine concentration range of 0.2-0.6 mg/L at the tap connections throughout the piped system. We selected this target free chlorine concentration based on a previous study which identified a median taste detection threshold of 0.7 mg/L from taste acceptability tests in Bangladesh (30). We also targeted this range because it aligned with the World Health Organization (WHO)’s recommendation for a minimum free chlorine residual in drinking water (0.2 mg/L) and when there is a known risk of fecal contamination (0.5 mg/L) (31). Following installation of the tablet dosers, Gram Vikas visited each village daily during the water supply time to iteratively adjust the dose control valves until the chlorine concentration was observed within the target range. Multiple days of testing and dose adjustment were necessary for controlling the chlorine concentration because water was supplied intermittently, which meant a dose adjustment on one day could only be assessed the following day when water was being supplied. Gram Vikas monitored the performance of the tablet dosers each week for the duration of the study by testing for free and total chlorine concentrations at 2 tap connections per village and providing refills and maintenance as needed. On average, Gram Vikas provided refills to the chlorinators every 21.4 days (SD = 12.6 days) depending on the village. Gram Vikas followed a protocol to proactively refill tablets before they were completely depleted; they provided refills when at least three partially dissolved tablets remained to prevent interruptions in chlorination.

After observing frequent no-dosing events, we increased the target chlorine concentration range to 0.4-0.6 mg/L in August 2024. By increasing the target chlorine concentration, we expected to observe improved disinfection and a greater proportion of tap connections with a detectable concentration of chlorine (> 0.1 mg/L). We also expected a decrease in no-dosing events caused by variability in the tablet dosers as chlorine tablets dissolved.

### Data Collection Procedures

Alongside our data collection partner, the Abdul Latif Jameel Poverty Action Lab (J-PAL) South Asia, we conducted baseline data collection in all study villages in October 2023 to understand usage of tap connections and water quality prior to implementing the in-line chlorination intervention. We conducted a census of all households with pregnant women or children under the age of 5 (N = 914) and asked questions of the female head of household related to demographics, WASH access, and health. We followed up with a subset of 200 households – who reported drinking water from their household tap connection – with a longer survey to characterize water treatment practices, perceptions of chlorination, and test for free and total chlorine residual at their tap connection. In a randomly selected subset of 80 households (out of 200), we also collected drinking water samples from tap connections and stored water containers and characterized microbiological contamination by quantifying total coliform and *E. coli* concentrations.

All surveys were conducted on electronic tablets using SurveyCTO, a computer-assisted personal interviewing software (Dobility, Cambridge, MA, USA). Drinking water samples were collected from household tap connections and water storage containers and tested for microbiological contamination using a defined-substrate assay, IDEXX, to enumerate the most probable number (MPN) of total coliform and *E. coli* per 100 mL of water (IDEXX Laboratories, Inc., Westbrook, ME, USA). Water samples were collected using sample collection bags containing sodium thiosulfate (Whirl-pak, Madison, WI, USA) to deactivate residual chlorine and transported to a field laboratory within 6 hours. Water samples were collected from tap connections after letting water flow from them at their highest flow rate for 30 seconds, after the water supply in the village had been on for at least 5 minutes. For stored water samples, respondents were asked to provide water from their storage container they would drink from. Samples were processed using the IDEXX quanti-tray system and incubated for 18-24 hours at 35 °C. Two trained lab technicians counted wells that were positive for total coliform and *E. coli* and a supervisor separately checked their counts. We took quality control measures to assess contamination including testing lab blanks on a daily basis, field blanks on a weekly basis, and duplicate samples in 2% of all samples. Field and lab blanks involved processing and analyzing separate 100 mL samples of sterile water collected either in the laboratory or the field. Free and total chlorine residual were tested using a N,N-diethyl-p-phenylenediamine (DPD) colorimetric method with HACH DR300 colorimeters (HACH, Loveland, CO, USA). We also tested for antibiotic resistant (ABR) *E. coli* in a random subset of samples from baseline and follow-up rounds using a modified validated IDEXX assay. The modified assay quantified phenotypic resistance of presumptive extended spectrum beta lactamase (ESBL)-producing *E. coli* by adding 80 µL of 5 mg/mL cefotaxime solution to each 100 mL sample (5,32). This resistance mechanism confers resistance to several critically important antibiotics, including third generation cephalosporins (5).

After implementation of the in-line chlorination intervention between December 2023 and January 2024, we conducted follow-up data collections until October 2024. In all 20 villages, 6 follow-up surveys were conducted in February, March, April, August, September, and October 2024. The monsoon season began in July 2024, which resulted in increased precipitation during the August, September, and October 2024 rounds. During the February, March, and April follow-up rounds, 10 households were randomly selected in each village (N = 200) from the baseline census list and surveyed about their water treatment practices, perceptions of chlorination, and had their tap and stored water tested for free and total chlorine residual. In a random subset of 4 households, we also collected drinking water samples from tap connections and stored water containers and quantified the MPN per 100 mL for total coliform and *E. coli* (N=80). In the August, September, and October follow-up rounds, we reduced the number of surveys to 4 randomly-selected households in each village due to budget constraints (N = 80). Testing for presumptive ESBL-producing *E. coli* was conducted during the October 2023, April 2024, and October 2024 rounds. An additional census of households with pregnant women and children under five was conducted in May 2024 to assess usage of the tap connections and adoption of chlorinated drinking water after being exposed to the intervention for 4 months (N = 886).

Independently from Gram Vikas, J-PAL South Asia visited each treatment village weekly, during which they tested drinking water from tap connections and stored water containers for free and total chlorine residual to assess if the concentration was within the target range. In the event a measured chlorine concentration was outside the target range, we informed Gram Vikas, who would take action to meet the target range either by adjusting the control settings or providing a refill. Tests were conducted at the nearest and farthest households in each village to assess if there was any significant decay in the chlorine concentration over the length of the distribution pipeline. We also tested for free chlorine at five minute intervals over the complete supply time at the nearest and farthest tap connections from the storage tank to assess any variation in dosing while water was being supplied.

### Outcomes and Statistical Analyses

Our primary outcomes included free and total chlorine residual and *E. coli* contamination in tap and stored water samples, reported usage of the piped water system, and water treatment practices. We created binary presence/absence variables for free and total chlorine residual with a positive detection threshold of 0.1 mg/L to account for the detection limit of the colorimeters and their error range. We created binary variables representing the presence or absence of total coliform and *E. coli* in tap and stored drinking water samples. We log_10_-transformed MPN estimates for *E. coli* and imputed non-detects with a value representing half the detection limit (0.5 MPN per 100 mL) to allow for log-transformation (33,34).

Prevalence ratios (PR) for all primary outcomes were estimated using Poisson regression. Counts of outcome observations were summed at the village-level and an offset covariate for the number of observations within each village was included in the model specification. Villages were followed up with over equal time periods. We also estimated average reductions in log_10_ MPN counts for total coliform and *E. coli* using linear regression applied to individual-level data with robust standard errors. All models included fixed effects for baseline outcome measurements for each village, to control for any imbalance between treatment and control groups at baseline. Analyses were performed using R version 4.3.1 (CRAN).

In March 2024, one treatment village requested their in-line chlorination device to be uninstalled following concerns about taste and smell of the drinking water. However, this village was still included in the analysis, thus estimating the intention-to-treat effect of our in-line chlorination intervention.

## Results

The baseline census identified 3,848 households in the 20 villages during October 2023. Among these, 914 households had children under five or pregnant women and participated in the census survey (N = 491 in control villages; N = 423 in treatment villages) (Table 1).

**Table 1:** Baseline Household Characteristics by Village-Level Treatment Assignment.

| Characteristic | N | Control | Treatment | Mean Difference (SE) |
| --- | --- | --- | --- | --- |
| **Panel A: Behavioral Outcomes** |  | **(N = 491)** | **(N = 423)** |  |
| Average household size | 914 | 5.69 (2.16) | 5.59 (2.00) | 0.10 (0.24) |
| Female head of household head gender | 914 | 24% (1.9) | 24% (2.1) | 0.0 (3.3) |
| Respondent can read and write | 914 | 55% (2.2) | 58% (2.4) | -2.8 (6.6) |
| Primary drinking water source | 914 |  |  |  |
| Household tap |  | 77% (1.9) | 81% (1.9) | -4.7 (7.3) |
| Borehole |  | 15% (1.6) | 9.5% (1.4) | 6.0 (5.3) |
| Community tap |  | 3.1% (0.8) | 7.3% (1.3) | -4.3 (2.4) |
| Covered dug well |  | 0.8% (0.4) | 0.5% (0.3) | 0.3 (0.8) |
| Surface water |  | 4.1% (0.9) | 1.4% (0.6) | 2.7 (2.6) |
| Uses a secondary drinking water source | 913 | 25% (2.0) | 33% (2.3) | -7.4 (9.2) |
| Drink water from their tap connection | 905 | 88% (1.5) | 92% (1.3) | -4.3 (4.0) |
| Women are primarily responsible for water collection | 200 | 78% (5.1) | 81% (2.8) | -3.0 (5.7) |
| Treats drinking water | 914 | 44% (2.2) | 48% (2.4) | -3.6 (6.6) |
| Time spent treating water > 5 minutes | 200 | 25% (4.3) | 37% (4.8) | -12.0 (11.4) |
| Women are primarily responsible for treatment | 85 | 92% (4.5) | 92% (3.3) | 0.6 (5.4) |
| Satisfied with taste of tap water | 199 | 97% (1.7) | 91% (2.9) | 6.0 (4.3) |
| Has electricity | 914 | 94% (1.1) | 96% (0.9) | -2.5 (1.7) |
| Owns a TV | 914 | 50% (2.3) | 56% (2.4) | -6.2 (5.9) |
| Owns a cell phone | 914 | 93% (1.2) | 94% (1.2) | -1.0 (2.2) |
| Owns a refrigerator | 914 | 23% (1.9) | 26% (2.1) | -2.3 (3.8) |
| Owns a motorbike or scooter | 914 | 49% (2.3) | 56% (2.4) | -6.1 (4.8) |
| **Panel B: Water Quality Outcomes** |  | **(N = 40)** | **(N = 40)** |  |
| Presence of total coliform at tap connections | 80 | 100% (0.0) | 100% (0.0) | 0.0 (0.0) |
| Mean log<sub>10</sub> MPN of total coliform at taps | 80 | 1.89 (0.66) | 2.15 (0.75) | -0.26 (0.26) |
| Presence of *E. coli* at tap connections | 80 | 70% (7.2) | 38% (7.7) | 32.5 (15.5) |
| Mean log<sub>10</sub> MPN of *E. coli* at tap connections | 80 | 0.32 (0.58) | -0.06 (0.42) | 0.39 (0.19) |
| Presence of total coliform in stored water | 80 | 100% (0.0) | 100% (0.0) | 0.0 (0.0) |
| Mean log<sub>10</sub> MPN of total coliform in stored water | 80 | 2.76 (0.75) | 2.68 (0.84) | 0.08 (0.20) |
| Presence of *E. coli* in stored water | 80 | 73% (7.1) | 60% (7.7) | 12.5 (14.0) |
| Mean log<sub>10</sub> MPN of *E. coli* in stored water | 80 | 0.81 (0.97) | 0.26 (0.79) | 0.55 (0.26) |

*Data are presented from the baseline census of 914 households and the longer baseline survey in a subset of 200 households. Averages (standard deviation) are reported for continuous outcomes, and proportions (standard error) are reported for binary outcomes. Mean differences*
*between control and treatment groups were estimated from linear regression models for each outcome of interest with standard errors clustered at the village-level.*

Most respondents reported using their household tap connection as their primary source of drinking water (77% of households in the control group, 81% of households in the treatment group). Respondents also reported using a secondary drinking water source to supplement their drinking water needs in 25% of control and 33% of treatment households. Treating drinking water before consumption was reported by 44% of control and 48% of treatment households. The reported water treatment methods used included boiling, filtering water through a coarse cloth, or allowing water to settle for a period of time before consumption. No respondents reported chlorinating their drinking water before consumption. Women were primarily responsible for water collection in 79.5% of households. In 19.5% of the households, both women and men were reported responsible for water collection, and in 1% men were responsible. In households that treat their drinking water, 92% reported that only women were responsible for treating water, 1% reported men were responsible for treating water, and 7% reported that both women and men were responsible. Respondents reported being satisfied with the taste of their tap water in 97% of control and 91% of treatment households.

We detected *E. coli* (>= 1 MPN/100 mL sample) in 38% of tap samples from treatment households and 70% of tap samples from control households (Figure 2). However, this baseline difference was not statistically significant after controlling for the block stratification and clustering standard errors for coefficient estimates at the village level (p-value = 0.14). No other differences in baseline measurements between control and treatment groups were statistically significant after controlling for block stratification. We detected *E. coli* in 60% and 73% of stored drinking water samples in the treatment and control households at baseline, respectively (Figure 3). Total coliform bacteria were detected in all samples from taps and stored water in both control and treatment households. At baseline, antibiotic resistant *E. coli* was detected in stored drinking water samples from 22% of control households and 15% of treatment households. We were unable to detect antibiotic resistant *E. coli* in any tap samples from control households at baseline, but detected it in 4% of tap samples from treatment villages.

**Figure 2:**
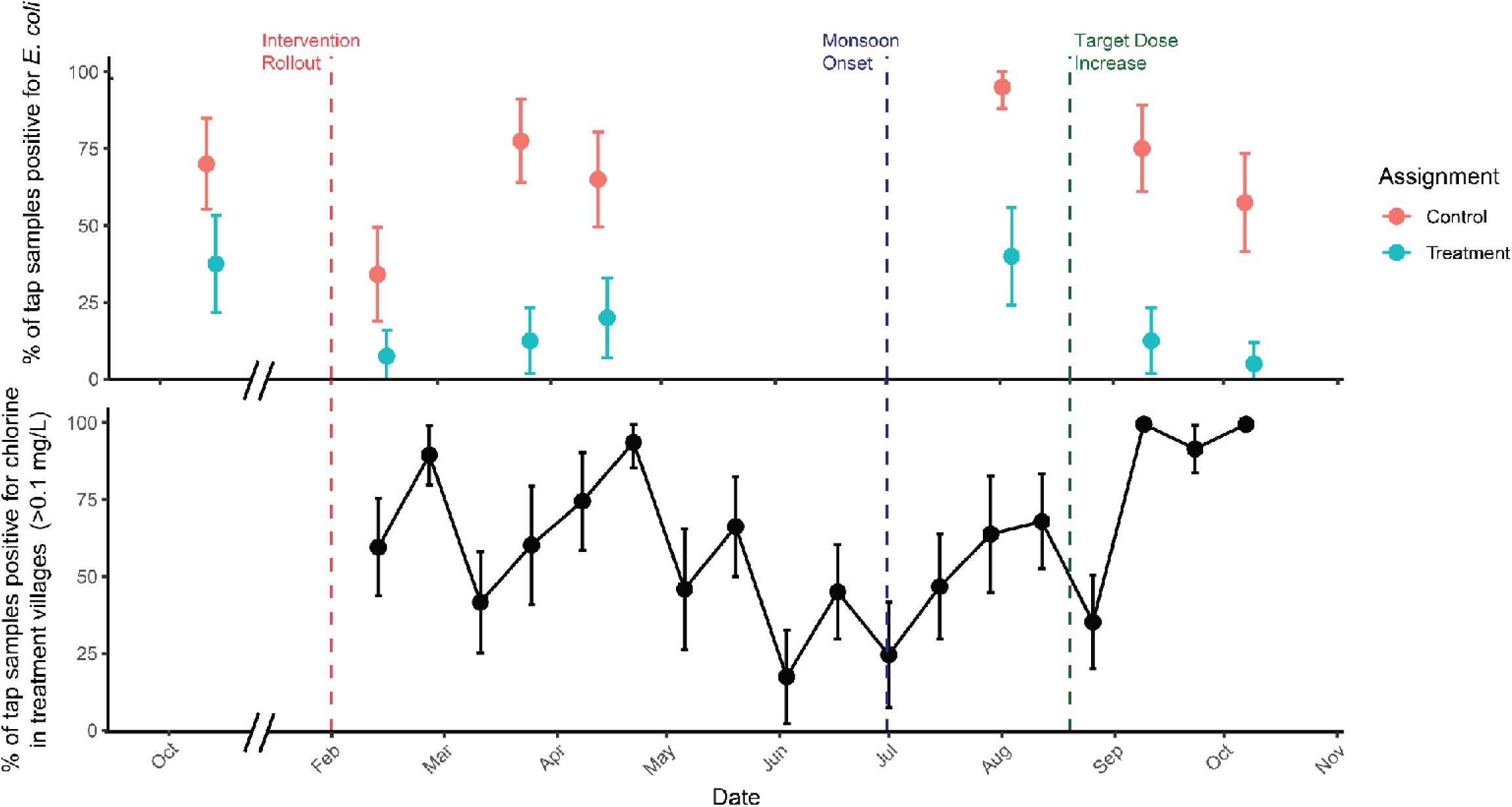
Comparison of *E. coli* Contamination and Free Chlorine Concentrations in Piped Water Supply by Treatment Status Over Time. Free chlorine concentrations were tested at 2 taps in each village on a weekly basis. The proportion of tap connections with detectable free chlorine concentrations (> 0.1 mg/L) were averaged across 9 treatment villages on a bi-weekly basis (N = 36 per time point). The averages reported above are unconditional means and do not control for randomization stratification variables. Although the difference in unconditional means at baseline is statistically significant (p-value = 0.03), the balance test controlling for stratification block and clustering standard errors at the village level is not (p-value = 0.14). One treatment village requested their chlorine doser to be uninstalled in March 2024, and we stopped weekly monitoring for free chlorine residual, but continued testing for E. coli contamination. The proportion of taps with a presence of E. coli was averaged across all 20 communities for each data collection round, stratified by treatment assignment. Error bars on both figures represent 95% confidence intervals. No confidence intervals around point estimates indicates that all water samples had free chlorine concentrations above 0.1 mg/L.

**Figure 3:**
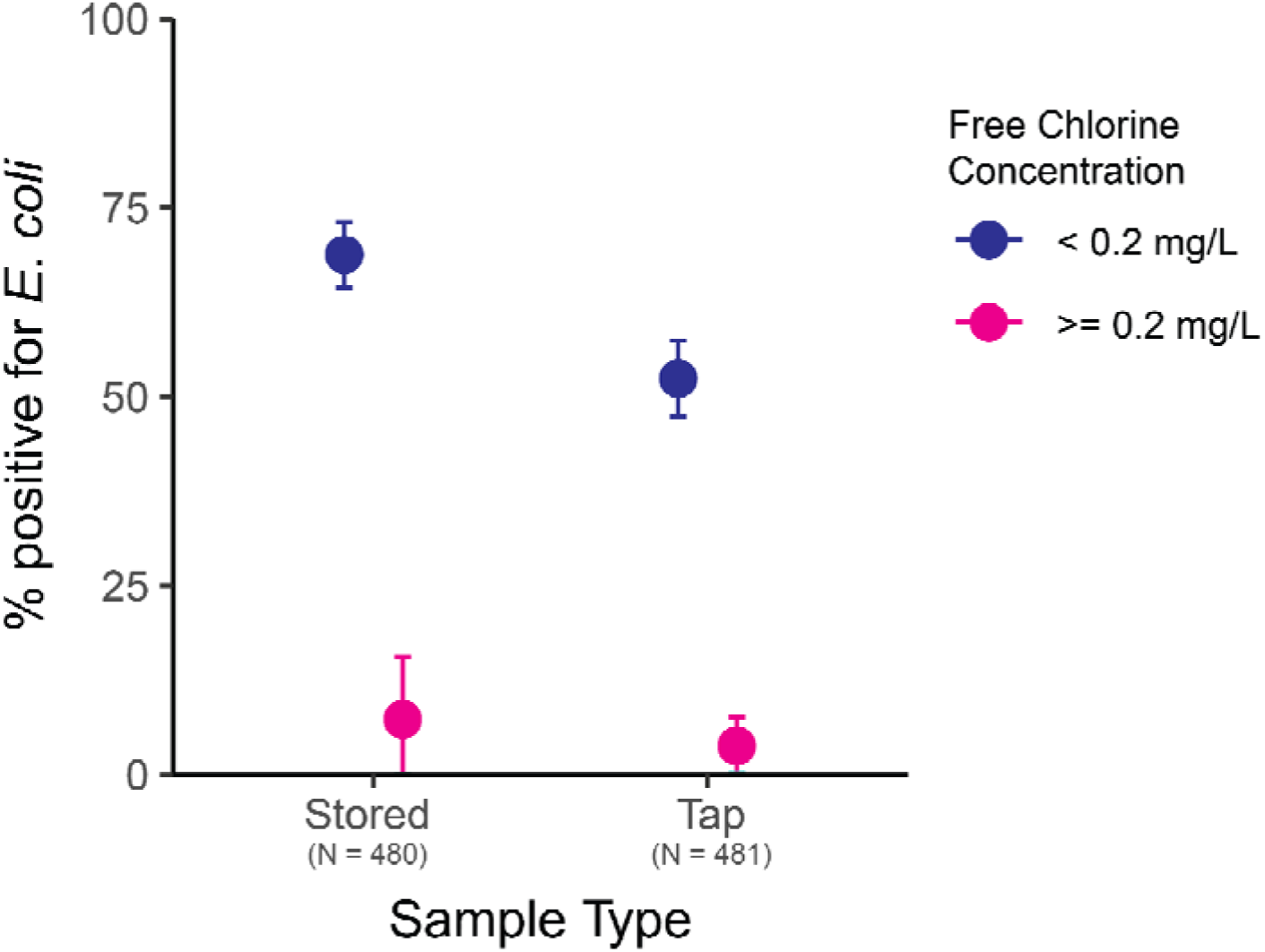
Comparison of *E. coli* Contamination and Free Chlorine Concentrations in Piped Water Supply Over All Follow Up Rounds. The proportion of tap and stored water samples with a presence of E. coli was averaged across all 20 communities for all 6 data collection rounds and stratified by their free chlorine concentration.

Over the study period, the proportion of tap connections with a detectable presence of free chlorine (> 0.1 mg/L) ranged between 18% to 100% during the weekly monitoring rounds. The low proportion of taps with chlorinated water would often occur after chlorine tablet refills were not provided in time or due to noncompliance by village pump/valve operators, who would turn off the chlorine dosers in response to taste and odor aversion by community members. At other times, the average chlorine dose was approximately 0.2 mg/L, and as chlorine tablets dissolved and their surface area and mass decreased, we hypothesized that the chlorine dosing rate decreased, and the average chlorine dose at tap connections would often drop below detectable concentrations. Following an increase in the target dose range to 0.4 to 0.6 mg/L in the 9 treatment villages still receiving chlorinated water in August 2024, the proportion of taps with a detectable concentration of free chlorine improved to between 92% and 100% during weekly monitoring.

Paired chlorine concentration measurements from the nearest and farthest tap connections differed by an average 0.11 mg/L (SD = 0.11 mg/L), indicating there was no major decay in chlorine concentration along the distribution network in any treatment village. The chlorine concentration at each tap connection was consistent across the full supply cycle in most treatment villages (SI Figure S2). In one village, a second borehole well was repaired in July 2024 and began to supply unchlorinated water to the central tank through an additional pipeline. The pump operator in the village would only operate this second borehole when water was being supplied to the village to extend the supply time, which meant the chlorinated water in the tank would become diluted with unchlorinated water from the second borehole. We observed a maximum decrease of 0.51 mg/L in the chlorine concentration at the taps in this village during the supply time.

A small proportion of water samples with a free chlorine concentration greater than 0.2 mg/L still tested positive for *E. coli* (Figure 3), which suggests those samples may have had inadequate contact time for disinfection (3.8% of tap and 7.3% of stored water samples with greater than 0.2 mg/L).

We detected chlorine (> 0.1 mg/L) in 51% of tap and 25% of stored water samples across 6 follow-up data collection rounds (Table S1). However, following an increase in the target chlorine dose to be 0.4 to 0.6 mg/L during the last data collection round, we observed an increase in the proportion of samples with chlorine to 80% of tap samples and 45% of stored water samples. Over 6 follow-up rounds, 67% of tap samples from control households tested positive for *E. coli* as compared to 16% of tap samples from treatment households (Figure 4). Similarly, 83% of stored water samples from control and 44% of samples from treatment households tested positive for *E. coli*. We observed ABR *E. coli* in 26% of stored water samples in control households and 8% of stored water samples in treatment households. We detected ABR *E. coli* in 11% of control tap water samples but not in any treatment samples.

**Figure 4:**
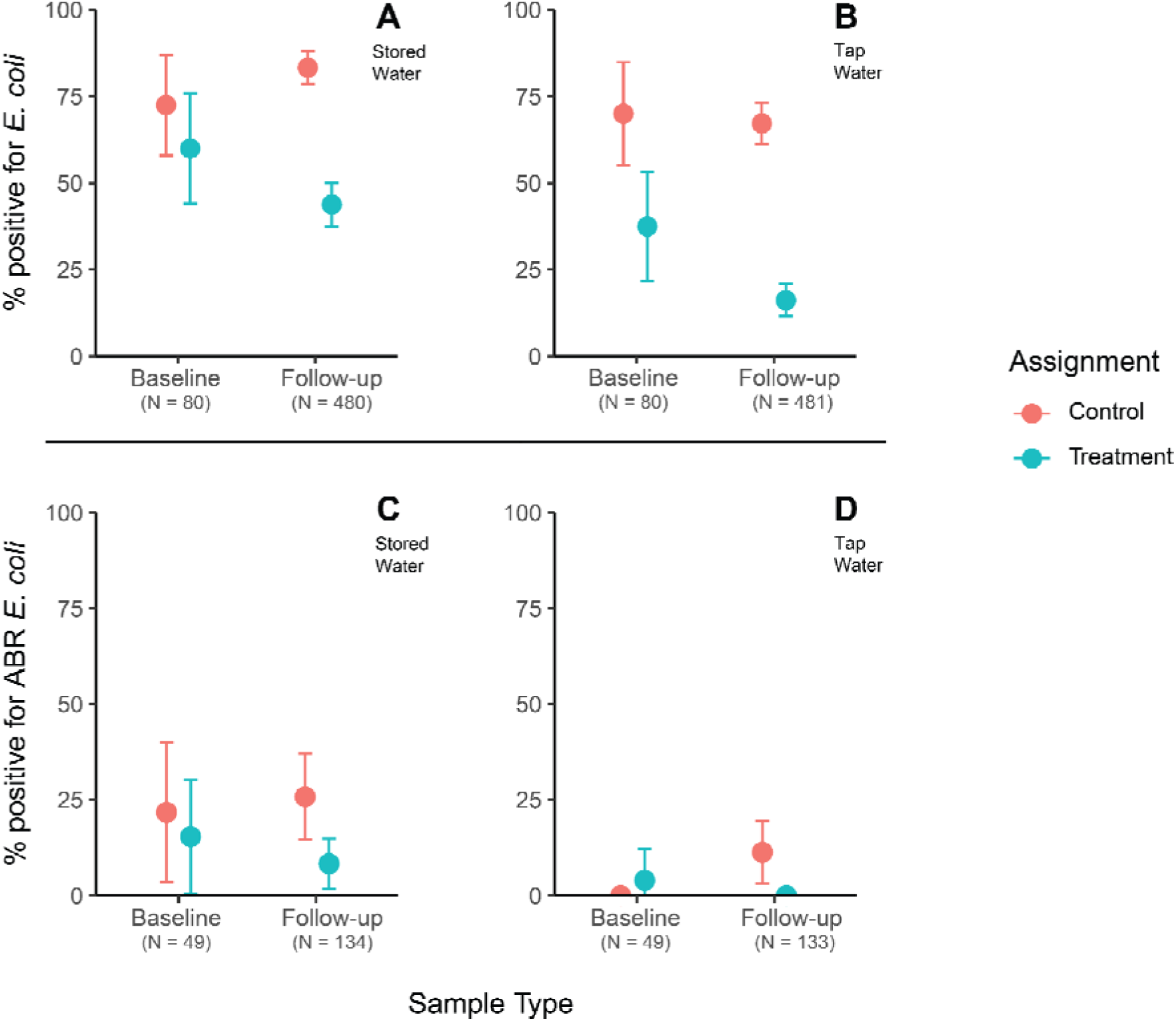
Comparison of *E. coli* Contamination By Treatment Status Over All Follow Up Rounds. The proportion of samples with a presence of E. coli was averaged across all 20 communities for stored water (Panel A) and tap water (Panel B) for baseline and all 6 data collection rounds and stratified by treatment assignment. The averages reported above are unconditional means. Antibiotic resistant E. coli contamination was also assessed during baseline and the April 2024 and October 2024 data collection rounds for stored water (Panel C) and tap water (Panel D). Error bars on both figures represent 95% confidence intervals.

After pooling microbial water quality data from all 6 data collection rounds, we estimate that treatment reduced the presence of *E. coli* in tap water by 70% as compared to the control group (PR: 0.30, 95% CI (0.20, 0.46); p-value < 0.001) (Figure 5). We also estimate that the presence of *E. coli* in stored drinking water was reduced by 47% in the treatment group as compared to the control group (PR: 0.53, (0.41, 0.68); p-value < 0.001). We observed an average reduction of 1.31 log_10_ MPN total coliform (□ = -1.31, 95% CI (-1.78, -0.84); p-value < 0.001) and 0.55 log_10_ MPN *E. coli* in tap water (□ = -0.55, 95% CI (-0.90, -0.20); p-value < 0.001) in treatment households as compared to control (Table 2).

**Figure 5.**
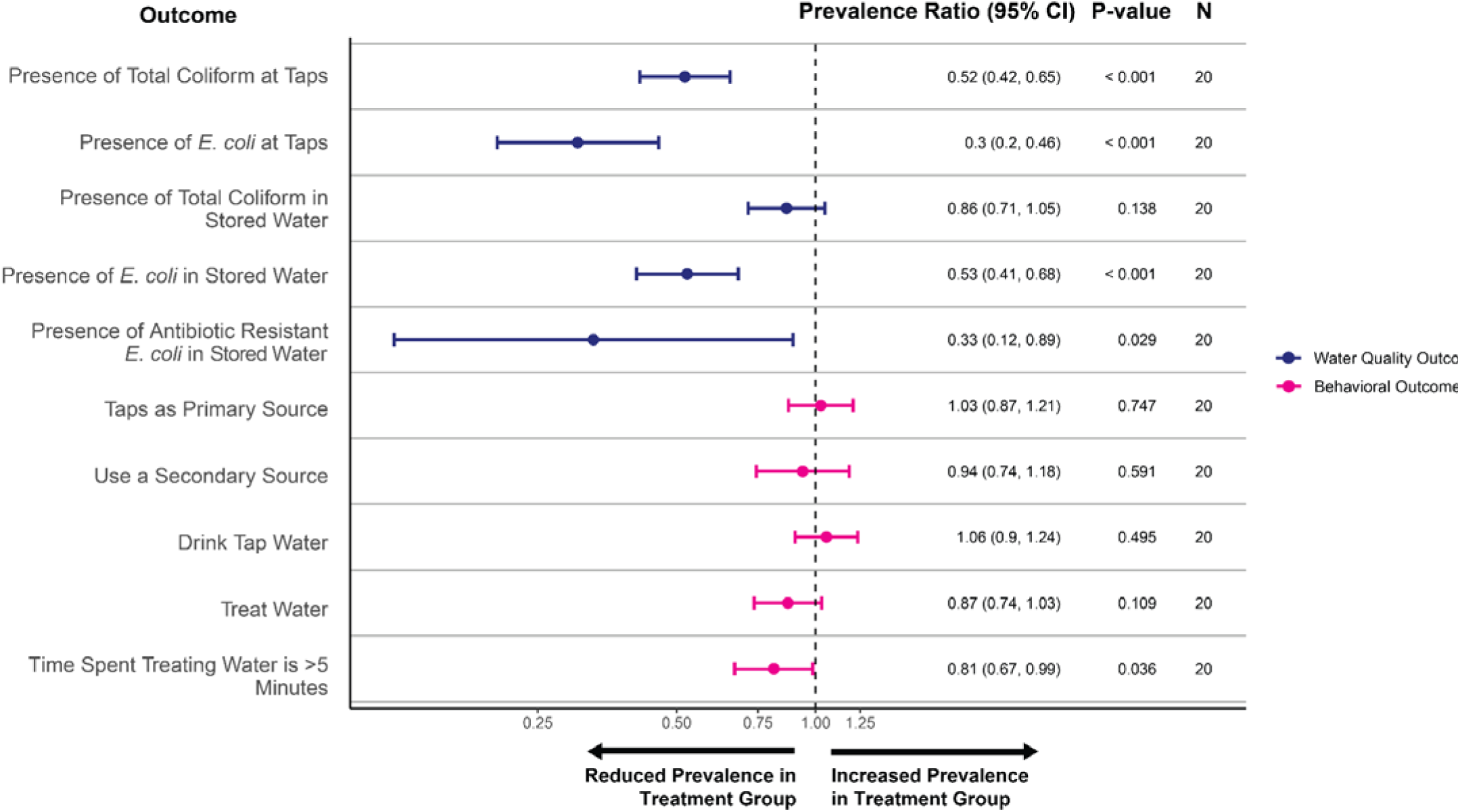
Treatment Effect on Key Water Quality and Behavioral Outcomes. Treatment effects on key water use outcomes were estimated after 4 months of exposure to the in-line chlorination intervention during a household census. Village-level prevalence ratios were estimated using Poisson regression with standard errors clustered at the village level. Model control for baseline measurements for each village. Linear regression results for the effects on log_10_ MPN E. coli and total coliform are presented in text only.

**Table 2:** Treatment Effect on Continuous Water Quality Outcomes.

| Characteristic | N | Control | Treatment | Difference (95% CI) | P-value |
| --- | --- | --- | --- | --- | --- |
| Mean log<sub>10</sub> MPN of total coliform at taps | 481 | 2.0 (0.9) | 0.6 (1.2) | -1.31 (-1.78, -0.84) | < 0.001 |
| Mean log<sub>10</sub> MPN of *E. coli* at tap connections | 481 | 0.6 (0.9) | -0.1 (0.7) | -0.55 (-0.90, -0.20) | < 0.001 |
| Mean log<sub>10</sub> MPN of total coliform in stored water | 481 | 2.7 (0.8) | 2.0 (1.4) | -0.74 (-1.04, -0.44) | < 0.001 |
| Mean log<sub>10</sub> MPN of *E. coli* in stored water | 481 | 1.0 (0.9) | 0.3 (1.0) | -0.63 (-0.90, -0.36) | < 0.001 |

*Differences were estimated from linear regression with standard errors clustered at the village level. All models control for the baseline average log<sub>10</sub> MPN of total coliform and *E. coli* in each village.*

We similarly observed a 0.74 log_10_ MPN total coliform ( = -0.74, 95% CI (-1.04, - 0.36); p-value < 0.001) and 0.63 log_10_ MPN *E. coli* reduction in stored water ( = -0.63, 95% CI (-0.90, -0.36); p-value < 0.001) among treatment households compared to control. We also estimated a reduction in ABR *E. coli* in stored drinking water following treatment (PR: 0.33, (0.12, 0.89); p-value = 0.29). We did not detect any ABR *E. coli* in tap water samples from treatment villages, but we detected ABR *E. coli* in 11% of control samples and therefore did not estimate a prevalence ratio for this comparison.

We did not observe any statistically significant differences between treatment and control households related to their use of household tap connections as the primary drinking water source (PR: 1.03, (0.87, 1.21); p-value = 0.747), use of a secondary drinking water source (PR: 0.94 (0.74, 1.18); p-value = 0.591), or consumption of tap water (PR: 1.06, (0.90, 1.24), p-value = 0.495). We saw a reduction in the proportion of participants who report treating their drinking water among the treatment group when compared to the control group (PR: 0.87, (0.74, 1.03); p-value = 0.109). We similarly observed a reduction in the proportion of participants spending more than 5 minutes treating their drinking water each day among the treatment group when compared to the control group (PR: 0.81, (0.67, 0.99); p-value = 0.036). Only 1 out of 475 participants (0.2%) in the control group mentioned an aversion to the taste or smell of water from their tap connection, as compared to 40 out of 405 participants (9.9%) in the treatment group.

## Discussion

Our study demonstrated that in-line chlorination, with dedicated monitoring, can effectively reduce fecal contamination in piped drinking water in rural India, without causing users to switch from the piped system to alternative, non-chlorinated drinking water sources. This is the first randomized implementation trial of in-line chlorination in India to demonstrate that chlorine tablet-based chlorinators could be adapted to operate under pressurized flow in piped water systems. Although we detected chlorine in only approximately half (51%) of tap samples during the first 6 data collection rounds in treatment households, this proportion greatly increased to 80% of taps during the final data collection round following an increase in the target free chlorine residual dose to a minimum of 0.4 mg/L. Previous evaluations of in-line chlorination involving chlorine tablet-based dosers in gravity-fed systems have detected a free chlorine residual in 40%-100% of water samples (17,35–39). It is difficult to isolate the reason for the variation across studies because flow conditions, distribution network lengths, target chlorine dosing ranges, and operational schemes vary both across and within studies. The results from this study and others highlight the importance of developing operational procedures to achieve the best performance of in-line chlorination, including the target chlorine concentration, management schemes, operator incentives, and refills frequency (37,38). Future in-line chlorination interventions can improve performance by targeting a free chlorine residual in the middle of the acceptable range (e.g. 0.5 mg/L to achieve a range of 0.2 mg/L to 0.8 mg/L) to account for fluctuating chlorine dosing due to chlorine demand in source water and variable performance of different in-line chlorination technologies. The acceptable range can be determined based on a local context’s water quality guidelines and taste and odor preferences, but the WHO recommends a minimum chlorine dose of 0.2 mg/L in drinking water (0.5 mg/L when there is a known risk of fecal contamination) (31).

We observed reductions in antibiotic resistant *E. coli* in drinking water among households with chlorinated water as compared to households with untreated water. Under the growing threat of antibiotic resistance transmission in the environment, the WHO’s Global Tricycle Surveillance project has recommended the use of ESBL-producing *E. coli* as a global indicator in environmental surveillance of antibiotic resistance (5). Drinking water has also been identified as an environmental reservoir of concern and recommendations have been made for the expansion of water treatment to limit antibiotic resistance transmission (7,9). Our study expands upon these recommendations by using presumptive ESBL-producing *E. coli* as a process indicator for controlling antibiotic resistance through in-line chlorination. Stored drinking water from treatment households was 67% less likely to have a presence of ESBL-producing *E. coli* as compared to control households, and we were unable to detect any ESBL-producing *E. coli* in treatment household tap samples during follow-up data collection rounds. Our study suggests in-line chlorination is an effective strategy for reducing human exposure to antibiotic resistant bacteria in drinking water. Future studies should consider the possible co-benefits of in-line chlorination in reducing diarrheal disease burden, mortality, and the burden of antibiotic resistant bacterial infections. Future studies could also consider adopting ESBL-producing *E. coli* as a water quality outcome to better understand how water treatment can reduce human exposure to antibiotic resistant bacteria.

Participants in treatment communities were less likely to spend greater than 5 minutes per day treating drinking water compared to participants in control communities. Few studies have quantified the time spent treating water associated with drinking water treatment interventions and who the responsibility falls to in the household (10,11,40). While other studies providing drinking water through continuous piped systems have estimated significant decreases in time spent collecting water (41,42), here we were able to document a reduction in time spent treating water as a result of having access to in-line chlorinators. One factor limiting take-up of point-of-use water treatment programs may be the time it requires for users (often women) to treat water (11). Our study provides evidence suggesting in-line chlorination can reduce the time burden associated with water treatment for individuals.

The introduction of in-line chlorination did not significantly change reported usage of government-provided taps for drinking water, despite reported aversion to taste and smell. Four months post-installation, households in treatment villages were just as likely to report using their tap connection as their primary source as control households. However, about 10% of respondents in the treatment group reported aversion to the taste and smell of chlorinated water. Respondents in treatment villages also anecdotally reported issues with cooking *pakhala*, a fermented rice meal that is a staple food of Odisha. Residual chlorine in drinking water disinfects microbial communities indiscriminately (43) and can alter the microbial communities responsible for fermentation in foods (44) and the gut microbiome in humans (45–47). The taste of chlorinated water has been cited as a barrier to adoption of chlorinated drinking water, particularly when users have to dose drinking water themselves (48,49). Median chlorine taste detection thresholds can also vary globally, though there have been limited studies (30,48,50–52). In the context of in-line chlorination, understanding taste detection and acceptability thresholds is important for setting a target chlorine dose which meets communities’ preferences to ensure high adoption. Lowering target chlorine concentrations in response to complaints without knowing if the complaints are associated with true aversion to taste and smell also presents a risk for adequate chlorination. When individuals can influence water treatment decisions in their community, designing the appropriate messaging and chlorine dosing procedures for new installations is important (30). While some have hypothesized that consuming chlorinated water over time increases acceptability thresholds (52), there have been no rigorous studies investigating if exposure over time affects the taste detection and acceptability thresholds of chlorinated water. Better understanding of how taste and odor acceptability changes with exposure to chlorinated water would be valuable for designing dosing strategies that maximize adoption, like, for example, starting at a low dose and increasing over some specified amount of time.

Participants in our study area used multiple water sources, and this practice affects exposure to non-chlorinated drinking water. Respondents would supplement their drinking water needs with water collected from unimproved water sources. Respondents commonly reported using alternative water sources when water was unavailable at their tap connection, which would occur due to intermittent water supply leading to limited availability or when groundwater could not be pumped due to electricity outages. Importantly, we did not observe increased use of alternative water sources after providing chlorinated water to households in the treatment group. Previous studies have highlighted convenience as being an important driver of water source selection (53,54), participants in our study may have continued use of their tap connection because it was conveniently located at their home. Consuming water from unimproved sources, even a few days out of the year, can significantly elevate exposure to fecal contamination and health risks (33,55). Having multiple water sources available for use can increase resilience to water supply disruptions caused by intermittent water supply, climate-related events, or humanitarian disasters and emergencies (53,56), but can significantly increase exposure to fecal contamination if those alternative sources are of lower quality. Not all secondary sources may be compatible with in-line chlorination and other water treatment methods may need to be considered for community-wide coverage in some settings.

Although the in-line chlorination devices effectively chlorinated water, we faced several operational challenges which led to inconsistent dosing or no dosing at all. First, although acceptance was generally high with only around 10% of respondents reporting issues with taste or smell, pump operators turned off devices at least once in all treatment villages in response to complaints. Gram Vikas responded to complaints by visiting and talking with village members, but this was not always successful at preventing pump operators from turning off the chlorinators. Community members in one village decided to have their chlorinator uninstalled because of taste and smell aversion following a community-wide meeting with Gram Vikas in March 2024. In another village, pump operators turned off the chlorinator after receiving repeated complaints about the taste and smell of the water from one household. Gram Vikas canvassed other households to understand the extent of complaints and learned that most households were in favor of the chlorinated water, but the pump operator still decided to turn off the device. Second, chlorine tablet deliveries to villages were occasionally delayed and led to no dosing events after refills were missed. Other studies have reported issues with consistent chlorine dosing including missed or delayed chlorine refills, clogging in the system due to precipitates from calcium hypochlorite tablets, damage to the water supply pipeline, design faults or broken parts, physio-chemical properties of water affecting tablet dissolution rates and chlorine demand, or decisions by communities to not purchase chlorine refills (36–39,57–59). It has been previously hypothesized that inconsistent chlorine dosing may reduce the likelihood that people become accustomed to drinking chlorinated water (17). In addition to social and behavioral communication and mechanisms to address complaints, future implementations of in-line chlorination should refine operational guidelines to address communities’ taste and smell preferences, and build the capacity of operators to ensure consistent chlorination.

These challenges highlight that achieving consistent chlorination will require community sensitization and systems to create incentives for workers to maintain and refill devices, and to keep them calibrated for the target dose. In our study, pump operators did not operate or maintain the chlorinators and we did not provide financial compensation to them based on guidance from the local Rural Water Supply and Sanitation Department. Pump operators therefore had limited incentives to keep devices switched on if some community members complained. During the study, we detected no-dosing events quickly due to regular monitoring of water quality. Delivery of in-line chlorination at scale would require similar monitoring to incentivize consistent delivery.

Other implementations of in-line chlorination have also achieved consistent chlorination in different infrastructure scenarios (37–39,58). Scaling in-line chlorination sustainably requires ongoing funding, as well as management systems which incentivize consistent refills and maintenance. However, there are several potential pathways to scale, including roles for both the public and private sector.

First, governments could decide to fund in-line chlorination. In October 2023, the Jal Jeevan Mission issued a national advisory on in-line chlorination adoption in single village schemes. After receiving preliminary results from this study and others (including pilots by NGO Tata Trusts in Assam and by Evidence Action in Madhya Pradesh and Andhra Pradesh) Jal Jeevan Mission published a handbook on water quality in piped water schemes in December 2024 recommending that “all states acquire chlorination systems” for rural piped water supply

(60). Some state governments have procured in-line chlorination devices at scale or initiated procurement. For example, Maharashtra has issued orders to construct over 17,000 electrochlorination plants (61). Chhattisgarh has issued an Expression of Interest seeking electrochlorinator manufacturers (62). Meghalaya has requested quotes for electrochlorinator installation and maintenance and is currently engaged in the bidding process (63). To our knowledge, several state governments have pilot tested tablet dosers and other ILC devices and are considering scaling up at the time of this writing (64). Odisha is also engaged in efforts to incorporate ILC into rural drinking water systems based on the results of this study.

Second, bundled payments for water treatment and water access could help to fund chlorination, as willingness to pay for water access is often higher than that of water treatment (16). Lindmark et al. (2023) evaluated a circuit-rider model for operating and maintaining in-line chlorination in rural Honduras and found that local government bodies charged tariffs for water access, part of which they used to purchase chlorine tablets from an NGO (58). The service provider’s salaries were supported by chlorine tablet sales, and their continuous service delivery led to more than 77% of water samples having greater than 0.2 mg/L over the last 9 years (58). Landlords and water kiosk owners operating community water systems have also demonstrated ability and willingness to pay for in-line chlorination devices and refills (65,66). In Bangladesh, a study documented that most landlords were initially willing and able to pay a monthly service fee covering the cost of installing and operating in-line chlorination devices (65). However, timely payments decreased over the service period, particularly for lower-income compounds. In Kenya, 27% of kiosk owners purchased an in-line chlorination device when offered the opportunity to buy (66). Six of these seven kiosks paid in full and 66% of their customers reported buying chlorinated water (they had the option to purchase from either non-chlorinated or chlorinated taps). Programs selling water treatment to intermediaries like kiosk owners or landlords could be supplemented with public subsidy.

Third, performance-based contracts with subsidies from governments or aid agencies could be used to incentivize utilities to use in-line chlorination by making payments conditional on reported water quality test results (67). In Malawi, the grant-funded organization Uptime made payments to a service provider based on the proportion of time hand pumps were reported functional, resulting in >95% uptime in the program area (65). At the end of 2024, Uptime reported 110,000 functioning water points with greater than 96% uptime across their global service areas in Africa, Asia, and Latin America with an annual payment of $0.43 per person served (68). In the context of in-line chlorination, water service providers could receive payments based on achieving a proportion of positive chlorine tests above a threshold at the point of collection in their water delivery system. Delivery models that incentivize consistent water treatment alongside water supply delivery are critical for scale up of in-line chlorination.

Our study has several limitations. First, the prevalence of fecal contamination was imbalanced at baseline in piped water between treatment and control villages, though this imbalance was not statistically significant after adjusting for block stratification and results were similar after controlling for baseline measurements in our models. Second, although we randomized the allocation of in-line chlorination, we did not randomize which villages received the PurAll 100 device versus the CTI-8. However, we did not observe any differences in performance between the technologies. Finally, we collected water samples after the water supply had been on for at least 5 minutes already, but in intermittent water supply schemes, the microbiological water quality is often at its worst during the first few minutes of supply because of a first flush effect (69), meaning we could have underestimated the magnitude of fecal contamination.

## Conclusion

This study suggests that in-line chlorination can improve water quality, lower the time burden of water treatment on individuals (usually women), and lead to increased and sustained user adoption of water treatment (11,14,15,19). Our findings also indicate in-line chlorination of drinking water should be considered as a strategy for reducing human exposure to antibiotic resistant bacteria. We recommend future in-line chlorination interventions set a target chlorine concentration in the middle of the acceptable range (e.g. 0.5 mg/L to achieve a range of 0.2 mg/L to 0.8 mg/L) for a given context to account for variability in the chlorine demand of source water and device dosing. To increase the scale and impact of in-line chlorination, operational procedures need to be developed for implementers to fit their context’s technical and social constraints.

## Acknowledgements

We sincerely thank the study participants for their involvement in this study. Thanks to the J-PAL South Asia team for their leadership in data collection including Archi Gupta, Archana M V, Nitin Kanakaraj, Prasanta Panda, and the entire data collection team. Thanks to the Gram Vikas team for their support in co-designing the intervention and implementing it, including Jobin Chacko, Varun Namineni, Debashis Mohapatra, Dibya Ranjan, Narottam Sethi, Kalicharan Panda, and Liby Johnson. Thanks to Luiza Andrade and Witold Więcek for advising and reviewing data analysis methods. Special thanks to Akanksha Saletore, Tavleen Singh, Shashank Patil, Michelle Cherian, Astha Vohra, and Niharika Bhagavatula for their leadership and support in conducting the study.

## Data Availability

All data and scripts are available on Open Science Framework https://osf.io/mx7z2/, DOI 10.17605/OSF.IO/MX7Z2

