## Supplemental Information for "In-line chlorination for drinking water in rural Odisha, India: a randomized controlled implementation trial"

**Supplementary Information**

Jeremy Lowe^1^, Vaishnavi Prathap^2^, Akito Kamei^3^, Sidhartha Giri^4^, Krushna Chandra Sahoo^4^, Michael Kremer^3^, Elisa M. Maffioli^5*^, Amy J. Pickering^1,6, 7*^

1. Department of Civil and Environmental Engineering, University of California Berkeley, Berkeley CA, USA
2. Development Innovation Lab - India, University of Chicago Trust, New Delhi DL, India
3. Department of Economics, University of Chicago, Chicago IL, USA
4. Indian Council of Medical Research – Regional Medical Research Centre, Bhubaneswar, Bhubaneswar OD, India
5. Department of Health Management and Policy, University of Michigan, Ann Arbor MI, USA
6. Chan Zuckerberg Biohub, San Francisco CA, USA

**Supplementary Information I: Additional details on primary study outcomes**

WASH variables were created using self-reported data on primary drinking water source use, usage of any secondary water sources, water treatment practices, time spent on treating water each day, and self-reported concerns about the taste and smell of the drinking water. Respondents were asked to recall all the drinking water sources they accessed in the previous month and the one they considered their primary source for drinking water. If respondents reported using their tap connection as their primary source or drinking water from their tap connection in the last month, we indicated this in a set of binary variables. If respondents reported using multiple drinking water sources in the previous month, we also coded this as using a secondary source. Respondents were also asked if they ever treated their drinking water before consumption and how much time it would take to treat their water, if at all. We created a binary variable indicating if a respondent spent more or less than 5 minutes treating their drinking water each day, the median time spent among respondents who treated their water. We also asked respondents if they had any issues with their drinking water, and, if so, what those issues were. From this, we created a binary variable indicating if a respondent had an issue with the taste and smell of their drinking water.

**Supplementary Information II: Baseline free and total chlorine testing**

The baseline presence of free chlorine in 6 tap and stored water samples are expected to be false positives given there were no reports of chlorinating drinking water. During baseline, colorimeters with a higher detection limit of 0.07 mg/L and less precise measurement (95% confidence interval of +/- 0.07) were used. These were replaced in subsequent data collection rounds with colorimeters with lower detection limits and more precise estimates.

**Supplementary Information III: Additional Figures and Tables**

**Table S1: Follow-up survey and census results**

| **Characteristic** | **N** | **Control** | **Treatment** |
| --- | --- | --- | --- |
| **Panel A: Behavioral Outcomes** |  | **N = 475** | **N = 405** |
| Primary drinking water source | 880 |  |  |
| Household tap |  | 74% | 76% |
| Community tap |  | 4.2% | 7.7% |
| Surface water |  | 0.8% | 0.2% |
| Borehole |  | 20% | 16% |
| Covered dug well |  | 0% | 0.2% |
| Other |  | 0.2% | 0% |
| Use a secondary drinking water source | 880 | 37% | 36% |
| Drink water from their tap connection | 880 | 79% | 84% |
| Treat drinking water | 880 | 76% | 66% |
| Time spent treating water is > 5 minutes | 880 | 60% | 51% |
| Reported smell or taste issue from tap water | 880 | 0.2% | 9.9% |
| Satisfied with taste of tap water | 599 | 99% | 94% |
| **Panel B: Water Quality Outcomes** |  |  |  |
| Presence of free chlorine at tap connection | 840 | 0.2% | 51% |
| Presence of free chlorine in stored water | 840 | 0.2% | 25% |
| Proportion of samples > 0.2 mg/L at tap connections | 840 | 0.2% | 39% |
| Proportion of samples > 0.2 mg/L in stored water | 840 | 0% | 18% |
| Proportion of samples > 2.0 mg/L at tap connections | 840 | 0% | 0.5% |
| Proportion of samples > 2.0 mg/L in stored water | 840 | 0% | 0% |
| Presence of total coliform at tap connections | 481 | 98% | 51% |
| Presence of *E. coli* at tap connections | 481 | 67% | 16% |
| Mean Log10 MPN of *E. coli* at tap connections | 481 | 0.61 (0.87) | -0.07 (0.66) |
| Presence of total coliform in stored water | 480 | 99% | 85% |
| Presence of *E. coli* in stored water | 480 | 83% | 44% |
| Mean Log10 MPN of *E. coli* in stored water | 480 | 0.98 (0.93) | 0.32 (0.95) |

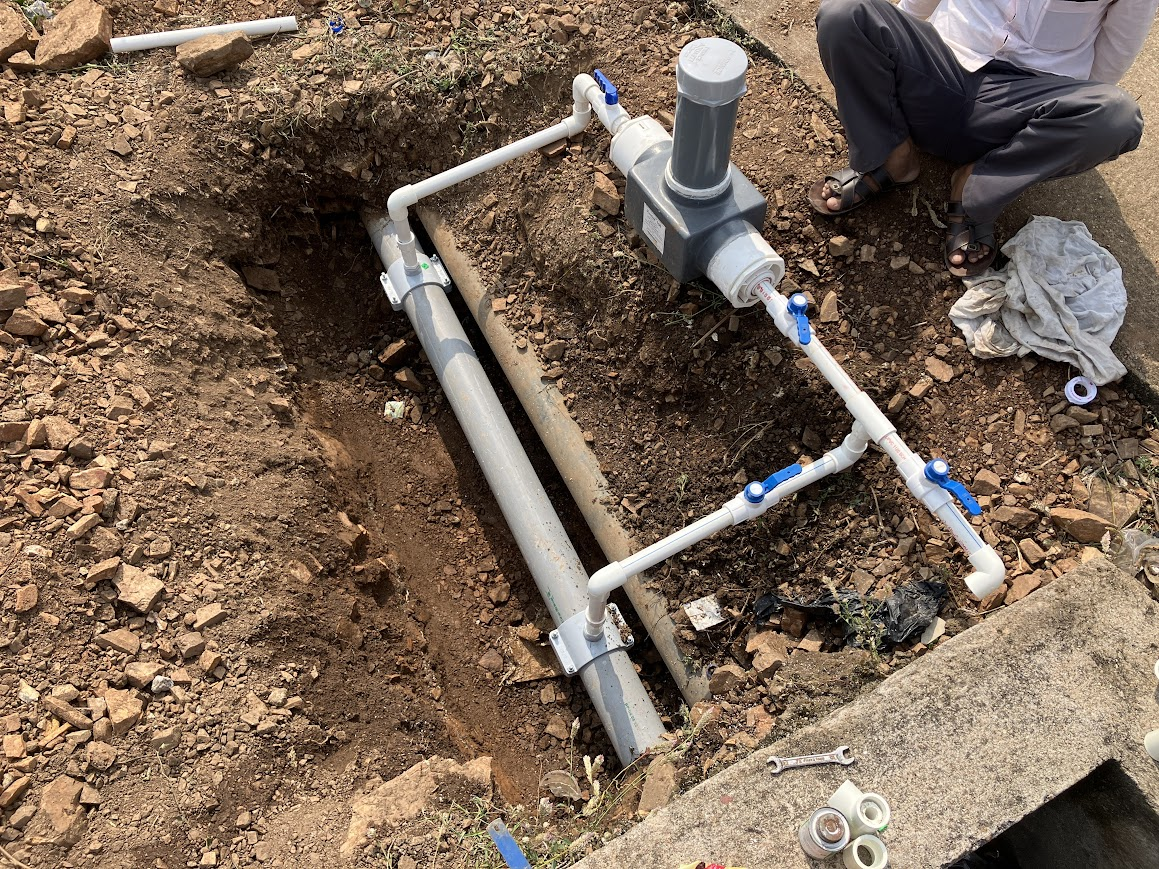

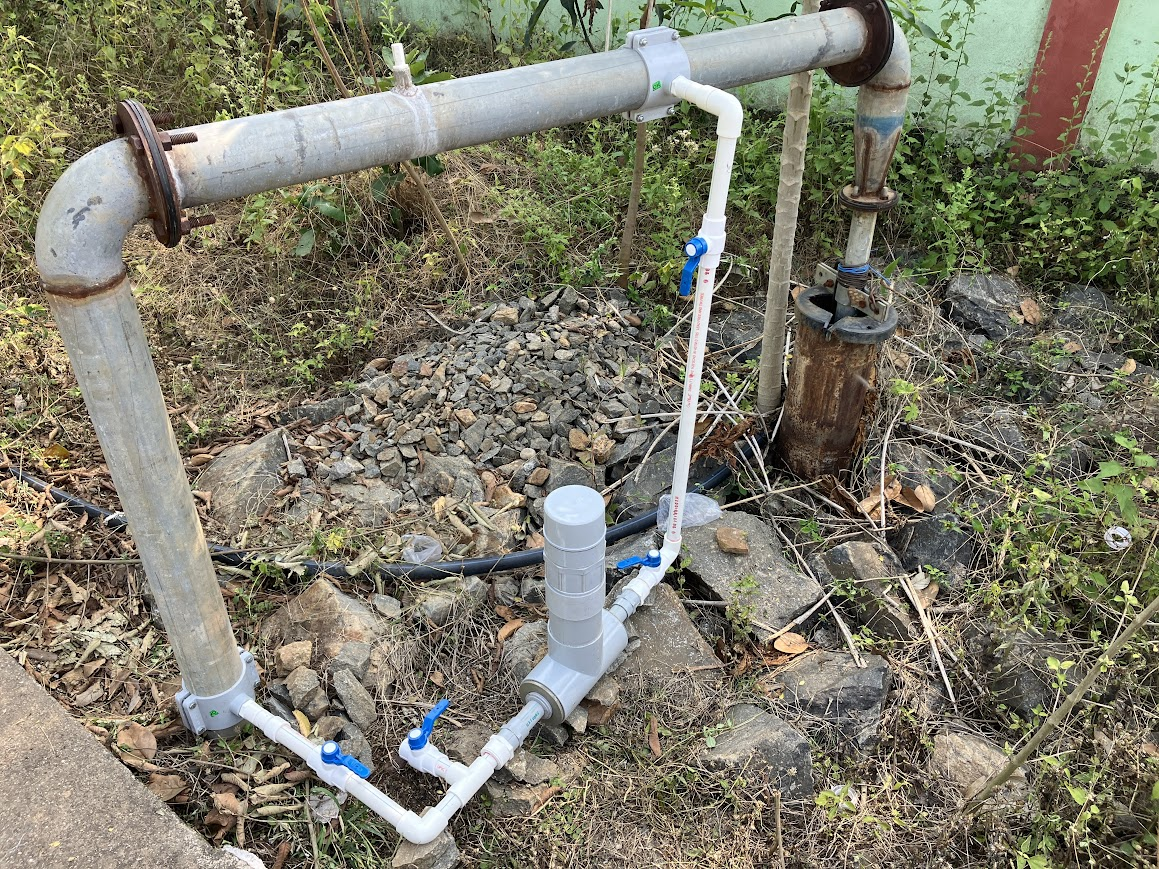

**Figure S1: Completed PurAll (left) and CTI-8 (right) ILC installations**

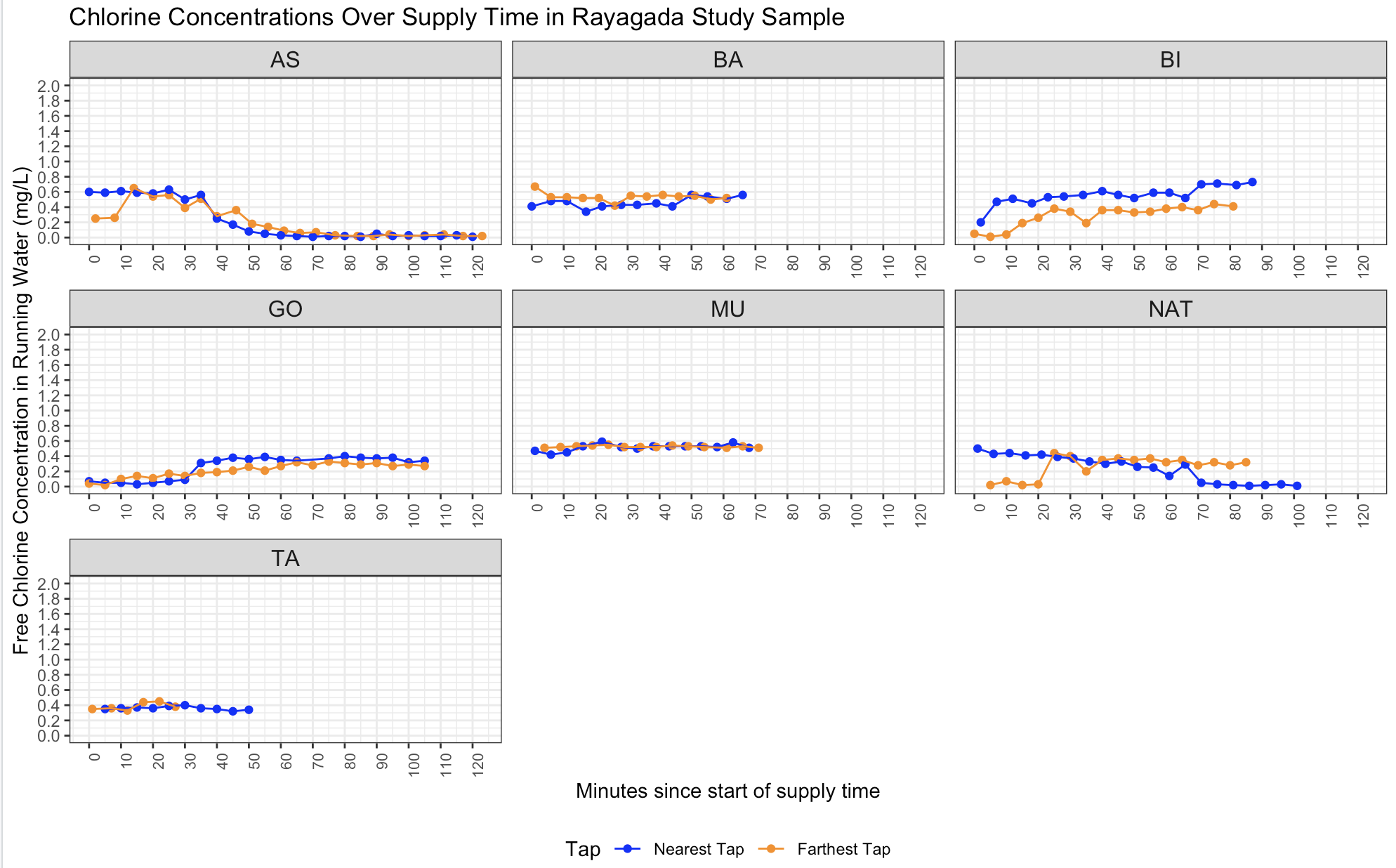

**Figure S2: Proportion of water samples with a presence of *E. coli* stratified by sample type and free chlorine concentration**
